# Aligning Artificial Intelligence Prediction Targets with Clinical Workflows Using Human Centered Design Methods

**DOI:** 10.64898/2026.01.15.26344209

**Authors:** Mark V. Mai, H. Stella Shin, Naveen Muthu, Nikolay P. Braykov, Andrea McCarter, Jordan Hilsman, Evan W. Orenstein, Xiao Hu, Swaminathan Kandaswamy

## Abstract

Artificial intelligence models in healthcare often fail to improve patient outcomes despite strong predictive performance because they are frequently developed with limited understanding of clinical workflows and system implementation. We demonstrate a human-centered design approach to define prediction targets before model development, ensuring alignment with actionable clinical interventions. Using pediatric acute kidney injury as a case study, we convened a multidisciplinary working group and applied three complementary methods: user stories to elicit role-specific prediction targets, a People, Environment, Technology, and Tasks (PETT) Scan to analyze sociotechnical system factors, and process mapping to identify workflow leverage points. This approach revealed that different clinical roles require distinct prediction targets, with shared barriers including inadequate monitoring practices, poor visibility of at-risk patients, and unclear trajectories for kidney injury progression. By integrating clinical context before algorithm development, we identified high-impact prediction targets that support actionable interventions for hospitalists, nephrologists, and intensivists, demonstrating how human-centered design can bridge technical model performance and real-world clinical utility.

## Introduction

Despite breakthroughs in computational power, data availability, and advanced algorithms, implementation of artificial intelligence (AI) models in healthcare still faces significant challenges and remains a work in progress. Of AI models developed for adults and children, only a fraction have described implementation.^1,2^ Similar to other healthcare interventions, these efforts have shown that AI models require careful integration and are not merely plug-and-play solutions.

To successfully integrate artificial intelligence at the bedside, developers must anticipate its fit within clinical workflows. Yet, AI models are commonly developed in technical isolation, a practice that often fails to account for the complex sociotechnical interactions and decision-making that define patient care. This paradigm has led to a well-documented disconnect between a model’s theoretical performance and its practical efficacy in improving outcomes - models that have been implemented often demonstrate strong predictive performance in silico, but in practice have failed to improve outcomes.^3^ To overcome this, collaboration between data scientists and clinicians is essential to develop the specifications for real-world use.^4^

In the automation research literature, experts have identified the key dimensions that one needs to account for design and implementation of an AI system.^5^ Here, we attempt to address two main areas: 1) what is the role of the proposed AI system versus the role of the clinician, and 2) which step within the human information processing pathway the AI system augments, how it fits into clinical workflows, and how alternatives are designed for performance trade-offs before implementation.^6^ Even when a diagnosis is suspected, various intermediate stages of a medical decision making process may need support from AI.^7^ For example, a system focused on 1) information acquisition may curate information from the EHR for the clinician to aid a task, 2) information analysis may synthesize patient data into an assessment about a patient’s current state, 3) decision and action selection may surface various treatment or monitoring options, or 4) action implementation may execute the actual action based on the recommended action.

In this work, we aim to explore how the specific step within human information processing supported by AI impacts selection of a prediction target, which is often overlooked prior to model development. Among these constraints, selection of the prediction target, integration into clinical workflow, and expected subsequent clinician action is arguably the most pivotal. When a prediction target does not align with an actionable intervention, the resulting model is unlikely to translate predictive power into better patient care.^8^ Similarly, AI must be designed with an understanding of the context of use by end-users. The core principle of human-centered design (HCD) is that technology must be shaped by the needs and workflows of its users.^9^ This provides a pragmatic framework to embed a deep understanding of clinical context – the problem, stakeholders, and workflow – directly into multiple stages of a model’s technological lifecycle. When applied to predictive modeling, this principle shifts the objective from achieving technical performance to enabling clinical action. Integrating HCD methods before development moves the focus from *what we can predict* to *what we should predict* to make a meaningful difference. The power of this approach lies in its focus on utility; even a model with average performance metrics can drive significant improvements in patient outcomes if it seamlessly integrates with and supports clinician decision-making.^6^ In this paper, we advocate for and describe a systematic methodology that operationalizes this principle, using HCD to define and prioritize prediction targets. We demonstrate its application through a case study focused on improving care for pediatric acute kidney injury (AKI).

Pediatric acute kidney injury (AKI) occurs when the nephrons that compose the kidney become damaged leading to impaired renal function, like issues with fluid retention electrolyte management, and acid-base buffering. AKI is a significant clinical challenge, affecting up to 55% of hospitalized children and leading to severe outcomes, including prolonged hospital stays and death.^10^ As a common condition associated with severe outcomes, AKI is a frequent target for predictive modeling, although none have been successfully implemented.^11–14^ Since there are no agents known to reverse AKI once diagnosed, AKI must be managed by preventing or mitigating risk factors – tasks deeply embedded in clinical workflow.^15,16^ To be effective, a model must trigger a specific, actionable intervention at the right time, which requires a deep understanding of the clinical environment. Therefore, in the pre-development stage for an AKI model, we applied HCD to systematically characterize the workflows and decision-making processes, ensuring the resulting prediction targets aligned with practical, high-impact interventions.

## Methods

We employed a three-phase approach to progressively refine our understanding of the clinical problem and identify the most impactful prediction targets: (1) user stories for initial target elicitation, (2) a People, Environment, Technology, and Tasks (PETT) Scan for sociotechnical system analysis, and (3) process mapping to identify leverage points – steps within the clinical workflow that would impact the largest number of patients.

### Setting

This study was conducted at a large tertiary pediatric health system with three free-standing children’s hospitals. This study was approved by the Children’s Healthcare of Atlanta IRB (STUDY00002289).

### Working Group

As part of a system-wide quality and safety priority initiative to reduce AKI rates in our healthcare system, our informatics team was tasked with performing exploratory data analyses and explore the potential for a predictive model to improve clinical care. To guide this effort, we convened a multidisciplinary working group focused on AKI. The working group met weekly consisting of clinical informaticists, data scientists, a human factors engineer, and clinicians (neonatology, nephrology, pediatric hospital medicine, and pediatric critical care medicine) (Table 1).

**Table 1.** AKI Working Group Members.

| Role | Working Group Members | Member Checking |
| --- | --- | --- |
| Pediatric Hospitalist | 2 | 1 |
| Pediatric Intensivist | 1 | 2 |
| Pediatric Nephrologist | 1 | 2 |
| Pharmacist | 0 | 1 |
| Data Scientist | 2 | 0 |
| Human Factors Engineer | 1 | 0 |
| Neonatologist | 1 | 0 |

### User Stories for Initial Prediction Target Elicitation

Rather than beginning with a predetermined prediction target based on data availability, we first sought to define what was clinically meaningful. We employed user stories as our primary method to capture the diverse perspectives of clinicians, ensuring that any potential model would be designed to answer a relevant clinical question and support a specific decision. To facilitate brainstorming of use cases and prediction targets, we adopted the following user story prompt format: “As a [persona], when I am [context], if we could reliably predict [prediction target], then we would do [intervention] which would lead to improvement in [outcome measure].”^17^ Through multiple iterations, we clarified the prediction targets for the various user groups at specific time points in their roles’ care processes.

### People, Environment, Tools, Tasks (PETT) Scan

To understand the complex interactions between clinicians (People), the physical care environment (Environment), existing technologies (Tools), and clinical workflows (Tasks), we performed a PETT Scan.^18^ This human factors method was chosen for its ability to deconstruct the work system and reveal hidden constraints or opportunities that a purely data-driven analysis would miss. During the weekly working groups, we conducted a PETT Scan to more fully consider the work system, factors to address, and intervention impacts on all work system factors, when identifying potential interventions that incorporate the potential prediction targets.^18^ Clinical members of the working group were each given a form for their roles containing a table listing their Goals, Facilitators (Environmental/Organizational, Process, Tools/Technology), and Barriers. Members were asked to complete these independently, considering their clinical roles, and then the forms were reviewed as a group, giving members an opportunity to amend their slides with additional considerations. We then performed member checking by presenting the data to additional clinical providers outside of the working group and asking them to add any additional items that they may consider (Table 1).

### Process Mapping to Identify Leverage Points

To synthesize our findings from the user stories and PETT scan, we constructed a clinical process map. This method allowed us to visualize the end-to-end patient journey in AKI care, integrate our understanding of system-level factors, and pinpoint the specific decision points where a predictive model could best support a suboptimal care process. Members of the working group integrated both clinical experience and the data from these two exercises to produce a process map, represented as swimlane diagram, covering from the beginning of a patient admission to various outcomes, such as discharge, need for renal replacement therapy, and death. The user stories were used to identify potentially suboptimal steps during decision making, which represented possible leverage points for an AI model. The process map and PETT scan were simultaneously reviewed by participants during working group meetings, followed by an additional member check, where clinicians who did not participate in the original working group reviewed the process map and the PETT scan (Table 1).

Finally, to prioritize the identified leverage points, we queried our EHR database to quantify the patient volume at each decision point. We included all pediatric inpatients admitted to the three Children’s Healthcare of Atlanta hospitals between August 1, 2018 and May 31, 2023, corresponding to our anticipated training dataset. This iterative, data-driven analysis of suboptimal decision-making steps allowed for data-driven prioritization based on potential clinical impact, which were reviewed by our working group.

## Data Availability

Data sharing is not applicable to this article as no datasets were generated or analyzed during the current study.

## Consent to Participate

Verbal consent was obtained from all participants included in the study.

## Clinical trial number

Not applicable

## Results

### User Stories

During our AKI working group, we constructed three physician user stories using the user story prompt, described in Methods. This exercise generated a candidate list of prediction targets, potential interventions, and outcome measures for provider-specific workflows (Table 2). The prediction targets for the 3 distinct personas included hospitalized patients at risk for AKI, AKI progression beyond KDIGO (Kidney Diseases Improving Global Outcomes) Stage 1, and AKI progression to Stage 3. During this exercise, participants generated the following possible downstream interventions: increased patient monitoring and lab screening, triaging rounding order, and adjusting clinical interventions (e.g. initiation of dialysis, fluid management). Outcome measures included AKI recognition, comprehensive AKI management, patients progressing to Stage 3 AKI, and time to AKI resolution. Notably, each user story was iterated upon multiple times after members of the working group considered the implications of the generated story. For example, for the pediatric hospitalist group, the prediction target was initially patients who would develop AKI. However, after realizing that most patients would recover from Stage 1 AKI, the target was refined to patients at risk of developing worsening AKI.

**Table 2.** Prediction target user stories table.

| User Role | User Story |
| --- | --- |
| *Example prompt* | *As a {persona}, when I am {context}, if we could reliably predict {prediction target}, then we would do {intervention}, which would lead to improvement in {outcome measure}.* |
| Pediatric hospitalist | As a hospitalist attending, when I am taking care of a patient that is at risk of developing worsening AKI, then we would increase patient monitoring and lab screening, which would lead to improvement in AKI recognition and action. |
| Pediatric nephrologist | As a nephrologist, when I am on the consult service in the hospital, if we could reliably predict which patients would not recover from Stage 1 AKI, then we would be able to prioritize our rounding order, which would lead to more timely and comprehensive management of AKI. |
| Pediatric intensivist | As an attending ICU provider, when I am taking care of a patient with Stage 2 AKI; if we could reliably predict that the patient would have worsening kidney injury leading to kidney failure, then we would *take appropriate clinical interventions* which would lead to improvement in *children reaching Stage 3 AKI or time to resolution of AKI*. |

### PETT Scan Findings

While these user stories provided initial, role-specific prediction targets, a PETT Scan was used to place these tasks within the broader clinical work system, revealing shared challenges and system-level barriers. The PETT scan resulted in a high-level characterization of the AKI clinical workflow (Table 3), organized into tasks (overarching user goals), barriers and facilitators (work system factors affecting tasks), people (individuals involved in the workflow), environments (physical and social contexts of tasks), tools (technologies and resources), and interactions (communication and coordination between people, tools, and environment).

**Table 3.** PETT Scan Tasks, Barriers, and Facilitators.

| Tasks | Recognition of AKI<sup>a,b,c,d</sup> |  | Reduce nephrotoxic medication exposure<sup>a,b,c,d</sup> |  | Identification of patients who would benefit from renal replacement therapy<sup>a,c</sup> |  | Monitor fluid status<sup>b,c</sup> |  |
| --- | --- | --- | --- | --- | --- | --- | --- | --- |
| Work System Factors | Barriers | Facilitators | Barriers | Facilitators | Barriers | Facilitators | Barriers | Facilitators |
| **People** | • Unnecessary nephrology consults, as most AKI will self-resolve<sup>a,b,c</sup><br>• Understaffed nephrology team<sup>a</sup><br>• Difficulty securing timely phlebotomist for lab draws<sup>a</sup><br>• Delayed nephrology consults for AKI can lead to suboptimal management, potentially avoiding dialysis<sup>c</sup> |  | • Absent or inaccessible pharmacist support<sup>a</sup><br>• PICU pharmacist shortage<sup>d</sup><br>• Staff pharmacist knowledge of drugs needing renal dosing<sup>d</sup><br>• Renal dosing tough without pharmacists<sup>a</sup><br>• Limited clinical pharmacy staff to safely review and manage all hospitalized patients<sup>c</sup> |  | • Limited CRRT trained ICU nurses<sup>b,c</sup><br>• Limited hemodialysis nursing staff – can only dialyze 2-3 hospitalized patients per day<sup>c</sup> |  |  |  |
| **Environments** | • False positive creatinine elevations in neonates<sup>a,b</sup><br>• Hospitalist culture deeming routine creatinine monitoring unnecessary<sup>a</sup><br>• Infrequent serum creatinine monitoring in at-risk patients leads to late AKI identification and missed early intervention opportunities, such as adjustment of medication dosing and fluid restriction<sup>c</sup> |  | • Polypharmacy decreases visibility of nephrotoxic medications<sup>b</sup> | • Pharmacy directed dosing of severe nephrotoxins<sup>b</sup><br>• Daily interdisciplinary rounds with pharmacists<sup>a</sup><br>• Pharmacy staff for consultation on drug adjustments, drug level monitoring<sup>c</sup><br>• New quality initiative NINJA improves attention to nephrotoxin exposure<sup>d</sup> |  | • CRRT usually rapidly deployed<sup>b</sup> | • Underlying conditions (e.g. distributive shock, ARDS, cancer) increase fluid overload risk with few modifiable factors.<sup>b</sup><br>• Interventions to decrease fluid overload, like diuresis, can induce AKI.<sup>b</sup> |  |
| **Tools** | • Poor visibility of AKI without opening charts<sup>d</sup><br>• Lack of in-house cystatin c testing misses AKI in some populations, like those with low muscle mass<sup>a,c</sup><br>• Unfamiliarity with NGAL and availability in-house<sup>b</sup><br>• Lack of built-in eGFR calculator to alert providers on low eGFR<sup>c</sup><br>• No diagnosis code for patients with a history of AKI/RRT, increasing risk of future AKI episodes (e.g., BMT, CHF patients)<sup>c</sup> | • Epic coloring of creatinine lab values<sup>a</sup><br>• Storyboard report for patients with reduced CrCl<sup>a</sup><br>• In-house urine NGAL availability, which may give some sense of whether the AKI is expected to resolve<sup>c</sup><br>• NINJA alert identifies patients at risk for AKI and recommends serum creatinine monitoring<sup>b,c</sup> | • High nephrotoxic drug exposure tool only appears for NINJA patients<sup>d</sup><br>• Poor perception of NINJA CDS<sup>b</sup><br>• NINJA alert does not alter management; only confirms clinical intuition<sup>b</sup><br>• Easy to miss nephrotoxins not in the hospital encounter MAR, like radiology contrast or meds given at an outside hospital prior to transfer<sup>c</sup> | • NINJA tools help clinical pharmacists identify high nephrotoxin exposure<sup>d</sup><br>• Column with color coded kidney designating adequate Cr monitoring for NINJA patients<sup>a,c,d</sup><br>• Reports with recent administration of nephrotoxins<sup>a</sup> |  |  |  |  |
| **Interactions between people, environments, tools and tasks** | • Unknown which patients with high AKI risk that will not rapidly reverse<sup>a</sup><br>• High prevalence causes trainees to ignore kidneys<sup>a</sup><br>• PICU transfers frequently uncommunicated AKI<sup>a</sup> |  | • Lack of medication adjustment when there is significant AKI - not just nephrotoxins, but antibiotics and others<sup>c</sup> | • Dosing guidelines and eGFR calculations help improve dosing<sup>d</sup> | • Delayed consulting of nephrology<br>• Unknown which patients would improve with CRRT and whether to start early<sup>b,c</sup> |  | • Culturally difficult to restrict fluids<sup>b</sup><br>• Nephrologists/pharmacists at times unwilling to adjust dialysate bags to support other organ failures<sup>b</sup> |  |

<sup>a</sup>Pediatric Hospitalists, <sup>b</sup>Pediatric Intensivists, <sup>c</sup>Pediatric Nephrologists, <sup>d</sup>Pharmacists

Several tasks were shared across different clinical roles, notably 1) recognition and identification of AKI, 2) reduction of nephrotoxic medication exposure, 3) identification of patients who would benefit from renal replacement therapy, and 4) monitoring of fluid status. Key barriers within the work system included:

- <u>People</u>: Understaffing among nephrology teams, phlebotomists, pharmacists, and dialysis trained nurses; knowledge gaps among pharmacy tech of drugs needing renal dosing
- <u>Environment</u>: A culture of de-emphasizing routine lab screening, prevalence of polypharmacy, and iatrogenic causes of AKI
- <u>Technology</u>: Poor perception of AKI and nephrotoxic CDS within the EHR, limitations of in-house cystatin c and urine NGAL (neutrophil gelatinase-associated lipocalin) testing, and the lack of a diagnosis code for patients with a history of AKI.
- <u>Workflow interactions</u>: Absent communication of AKI status during patient transfers, lack of knowledge of which AKI cases will reverse and which will worsen, and difficulties in coordinating care between nephrologists, pharmacists, and other clinicians.

Facilitators were identified less readily, but included the valuable role of pharmacists during rounds and for drug consultations, rapidly deployable dialysis resources, EHR color coding for creatinine lab values, a pre-existing alert system for identifying patients at risk for kidney injury (Nephrotoxic Injury Negated by Just-in time Action [NINJA]^15^), in-house urine NGAL testing, and dosing guidelines to help reduce nephrotoxic medication exposure.

### Swimlane Diagram and Workffow Analysis

Integrating findings from the user stories and the PETT scan, we created a swimlane diagram (Figure 1) that illustrates a patient’s hospital course from admission through determination of AKI risk, risk mitigation steps, and adverse kidney outcomes across various roles on the care team. These findings gave us view of the tasks and gaps where interventions could potentially be introduced to improve clinical care. The process map (Figure 1) was pivotal in shifting the focus from a single prediction point (initial AKI risk) to the entire care continuum. It visually demonstrated that the critical clinical uncertainty was not *if* AKI would occur, but rather *which* patients with early AKI would progress to more severe stages – highlighting a priority prediction target identified by pediatric hospitalists (Table 2). Furthermore, later along the course of worsening AKI, the process map helped visualize the challenges surrounding cases of severe AKI that do not respond to medical management.

**Figure 1.**
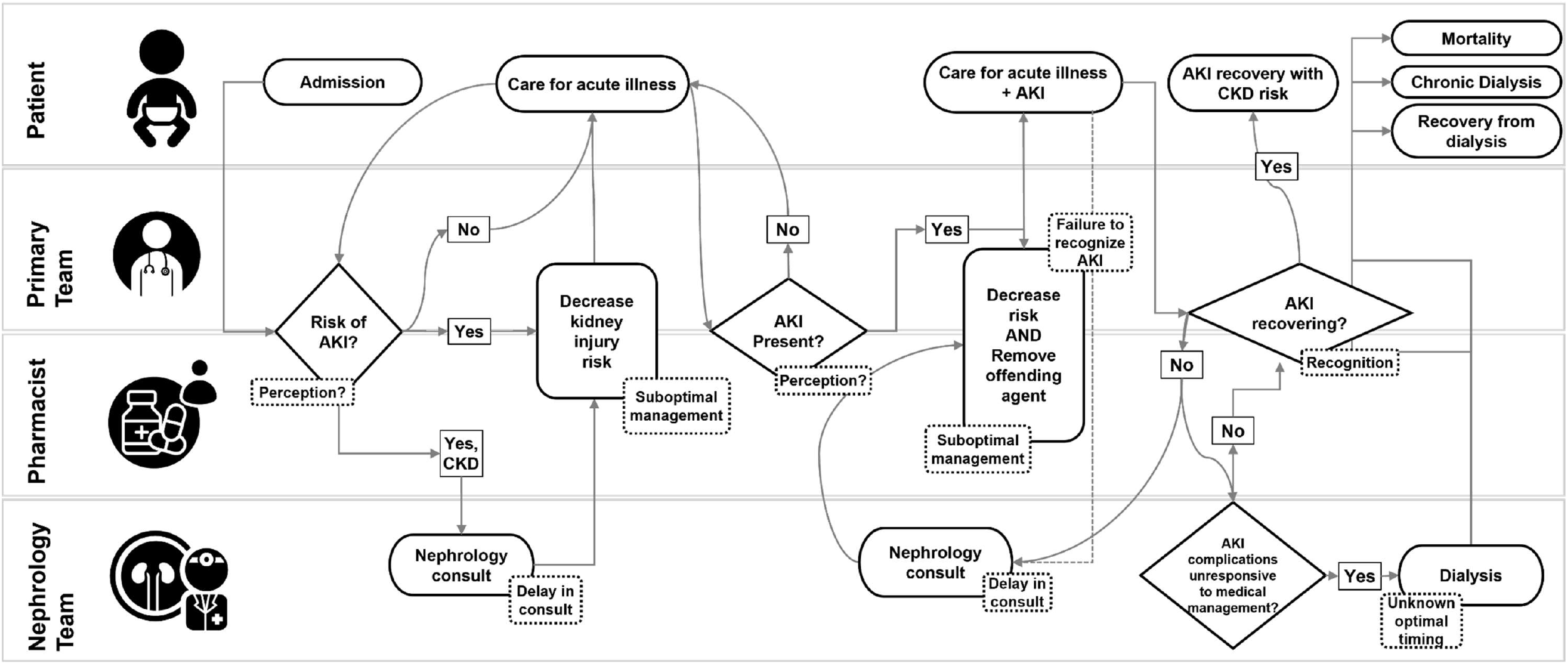
A swimlane diagram maps the decision points along a clinical workflow potentially amenable to predictive models. The flowchart depicts decision points (diamonds) and care processes (ovals) from hospital admission through final outcomes. Dotted boxes indicate potential care gaps including delayed recognition, suboptimal management, and consultation delays. The pathway shows role-specific responsibilities across patient care teams, pharmacists, and nephrology consultants, with branches leading to mortality, chronic dialysis, AKI recovery with CKD risk, or dialysis recovery. Key intervention points include nephrology consultation triggers, medication adjustments, and recognition checkpoints distributed across primary, pharmacy, and nephrology teams.

### Quantification of Leverage Points

Our analysis yielded three competing prediction targets: (1) incidence of any AKI amongst all hospitalized patients (high volume, low actionability), (2) amongst patients with AKI, patients who are likely to progress to more severe stages (moderate volume, high actionability), and (3) established AKI patients who are at risk of becoming unresponsive to medical management with progression to severe complications (low volume, high clinical impact).

The first target – predicting AKI incidence among all hospitalized patients – would address a large population (10% prevalence; n = 39,286/371,954 encounters). However, HCD principles revealed critical misalignment between predictive capability and actionable intervention. The majority of cases resolve spontaneously, and an existing clinical decision support (CDS) tool already mitigates a primary risk factor. The disconnect between prediction and intervention exemplifies how high population coverage does not guarantee clinical utility – a model accurately identifying patients who require no intervention provides limited value to care teams.

The second target – predicting AKI progression among patients with established disease – emerged as more aligned with clinical workflow and actionable decision-making. In our cohort, 17% of encounters demonstrated disease escalation (n = 3,559/21,409), with over 50% progressing from stage 1 to stage 2. This target offered two key advantages through the HCD lens: it enriches the population for targeted CDS integration, and it supports proactive clinical intervention before disease becomes refractory to medical management. By focusing on patients already identified with AKI, the model integrates naturally into existing surveillance workflows while enabling clinicians to intensify monitoring and treatment at a stage where interventions remain effective.

The third target – predicting progression to severe complications – addressed the most clinically compelling problem. Among patients with established AKI, 16.8% met a composite outcome of death, persistent renal dysfunction, or dialysis initiation, representing a population where prediction could meaningfully impact outcomes. However, the absolute number of patients was substantially smaller, limiting the model’s potential reach and the feasibility of developing robust predictive performance.

### Aligning Prediction Target with Clinical Context

Guided by HCD principles that prioritize utility over technical performance alone, we selected the second target: predicting which patients with established AKI would progress to worse stages. This decision reflects the core tenet that technology must be shaped by clinical needs and workflows. The target balances sufficient population size to support model development with clear opportunities for actionable intervention. Existing therapeutic approaches – optimizing fluid management, medication adjustments, and intensified monitoring – can be more effectively deployed when clinicians receive early warning of likely progression. This alignment between what the model predicts and what clinicians can act upon positions the model to translate predictive performance into improved patient care.

## Discussion

Human-centered design approaches clinical workflows proved essential for initiating development of clinical actionable AI prediction models, fundamentally shifting how prediction targets are defined and prioritized. Using pediatric AKI as a case study, our structured approach using standardized use case templates enabled clinicians to explicitly link prediction targets with specific downstream interventions, resulting in prediction model specifications to our data science team rooted in actionable clinical tasks. We also found that contextualizing these tasks within the broader workflow for AKI helped assess the potential patient impact and identify leverage points, which informed model development prioritization. With this approach, we identified a novel prediction target that would have been missed if we had taken the traditional approach of solely predicting patients at risk of AKI. Currently, there are no published pediatric prediction models progression of AKI stage.

Our findings demonstrate how HCD methods reveal that different prediction targets correspond to distinct stages of human information processing. The hospitalist prediction target (identifying patients at risk of worsening AKI) primarily supports information acquisition by curating which patients warrant increased surveillance and laboratory screening. The nephrologist target (predicting non-recovery from Stage 1 AKI) serves information analysis by synthesizing longitudinal patient data into trajectory assessments that inform consultation prioritization. Finally, the intensivist target (predicting progression to Stage 3 AKI) supports decision and action selection by surfacing when to initiate specific interventions like renal replacement therapy or aggressive fluid management. Notably, none of the identified use cases required action implementation – autonomous execution of interventions – highlighting that current clinical workflows favor augmentation of human decision-making rather than automation.

Current clinical AI development prioritizes model performance metrics over workflow integration, creating a critical gap between technical capability and clinical utility. Much of the lost promise of AI systems is likely due to malalignment of a model and its designed prediction task with work as done at the bedside. In fact, our team has previously found that while high-performing models will have degraded model performance metrics following implementation, careful integration with clinical workflows can still yield improvement in patient outcomes.^6^ This observation aligns with automation research showing that the stage of human information processing targeted by AI fundamentally shapes its clinical utility.^5^ A model supporting information acquisition (e.g., surfacing at-risk patients) has different performance requirements and workflow integration needs than one supporting decision selection (e.g., recommending specific interventions). By explicitly mapping prediction targets to information processing stages during the pre-development phase, we ensured that model specifications matched the cognitive support clinicians actually needed.

Despite growing recognition that AI systems must be embedded within sociotechnical frameworks, no systematic methodologies exist for aligning AI models with actual clinical workflows during the development phase. While performance metrics remain the primary benchmarks for model comparison and development progress – being readily computable and allowing straightforward ranking – they inadequately capture the critical dimension of workflow integration. Quantifying the degree to which models fit into clinical workflows represents an essential but underdeveloped area of inquiry. Lindsell et al^8^ advocate for starting AI development with proactively asking what clinical outcomes the AI tool will address, requiring defining the characteristics of affected patients and clinical settings, as well as how and to whom the results will be provided. McCradden et al^19^ elaborate on “action-informed” AI described by Lindsell et al^8^ noting that clinical relevance of a model is often distilled to model accuracy on a prediction problem that clinicians feel is important, but may in actuality be divorced from downstream potential clinical actions that benefit patients. Instead, they argue that AI systems should be situated within a larger intervention ensemble that recognizes that AI systems require a supporting structure of practices, both social and technical in nature, around them to effectively translate models into practice. Our study shows that there may be missed opportunities for prediction target selection without careful consideration of users and their clinical workflows.

Our work addresses this gap by operationalizing key components of “action-informed” AI development and “intervention ensemble’” framework.^8,19^ While these authors advocate for considering downstream clinical actions during AI development, we provide concrete human factors tools to systematically elicit and specify these actions, particularly focusing on use case definition, task specification, and outcome measurement. We iteratively used tools for human-centered design to more clearly define the problem being addressed with a predictive model, from prediction targets to workflow decision points that represent modifiable gaps in the care process. The user story framework forced consideration of which stage of information processing required support - whether clinicians needed help identifying relevant data (acquisition), interpreting patient trajectories (analysis), or choosing among intervention options (decision selection). This granularity prevented the common pitfall of developing a model that answers the wrong question at the wrong decision point.

The findings described here provide a practical roadmap for cross-functional teams of clinicians, informaticists and data scientists that are undertaking predictive modeling projects. We recommend that teams formally dedicate a ‘pre-development’ phase focused on workflow analysis and stakeholder engagement. This initial investment in understanding the clinical context, using methods like user stories and process mapping, can prevent the costly development of technically sound but clinically irrelevant models. Shifting the focus from ‘Can we predict X?’ to ‘If we could predict X, what would we do differently, and what would the impact be?’ is the foundational change for which our work advocates.

Beyond pediatric AKI, this structured human-centered design process – iteratively combining user stories, sociotechnical work system analyses, and process mapping – offers a reusable template for defining prediction targets that are anchored in real clinical decisions rather than data availability alone. By explicitly surfacing who is involved in decisions, where information is missing, and which steps in the care pathway are most modifiable, these methods can be applied to other prediction problems such as sepsis, emergent deterioration, or chronic disease management to identify targets that are both clinically meaningful and technically feasible. Extending this approach to explicitly examine which patients are systematically under-represented in workflows, monitoring practices, or digital touchpoints also creates an opportunity to embed digital health equity considerations upstream in model design, for example by making visible patients who are less frequently monitored, less likely to trigger existing alerts, or less likely to access digital tools. In this way, human-centered design can function not only as a tool for improving workflow fit, but also as a pragmatic lens for identifying and mitigating ways in which predictive models might otherwise perpetuate or exacerbate inequities in access and outcomes.

Several limitations warrant consideration when interpreting these findings. The single-institution design limits direct generalizability of specific workflow patterns and use cases, though the human factors methodology itself should transfer across settings. While we have identified prediction targets, we have not assessed other potential design opportunities, like identifying roles for the clinician and the AI in an AI-augmented workflow, where and how the model should be integrated into the workflow, and thinking of the impact on human performance. These will be the foci for future studies. The specific prediction target identified is tailored to our institution’s specific workflows and resources; other institutions using this methodology may arrive at different but equally valid targets. More importantly, our focus on AKI – a condition with relatively well-defined clinical pathways – may not reflect the complexity of applying these methods to more ambiguous clinical scenarios. The study’s pragmatic mixed methods, while appropriate for method development, lacks the rigor of formal qualitative research methods. The current study does not consider the role of AI systems compared to clinicians, nor does it assess integration into clinical workflows and the range of user interactions that may flow from that. Future study will be focused on this. Future applications would benefit from structured qualitative analysis to identify patterns across different clinical domains and organizational contexts. We also did not interview all inpatient hospital services that might care for patients with AKI, but instead focused on those that deal with AKI most frequently during inpatient stays.

Future directions for this work include the eventual implementation of AKI models developed using this process, their CDS development, and their impact on clinical care. Our team also is employing this human factors-based approach towards other use cases, including sepsis and emergent deterioration. In addition, other methods from human factors may be helpful in creating boundaries for AI system features, like performance threshold setting. We also plan to perform usability testing to elucidate which AI-driven clinical decision support components are most important to users embedded into their clinical workflows.

## Conclusion

HFE methods used prior to model development can clarify prediction targets and identify workflow enhancements and model success criteria that inform model development and deployment approaches.

## Author contributions

M.V.M., S.K., E.W.O., and N.M. conceptualized the study. M.V.M. and S.K. wrote the main manuscript text. M.V.M., S.K., E.W.O., and N.M. developed the methodology. N.B., A.M., and J.H. performed data curation and quantitative analysis of leverage points. M.V.M. and S.S. conducted validation. N.M., S.S., N.B., and X.H. provided critical revisions to the content. All authors contributed to writing - review & editing. S.K., X.H., and E.W.O. provided supervision. All authors read and approved the final manuscript.

## Acknowledgements

Mark V Mai’s work was supported by the National Center for Advancing Translational Sciences (NCATS) of the National Institutes of Health (NIH) under Award Numbers UL1TR002378 and KL2TR002381. Swaminathan Kandaswamy’s work was supported by the Agency for Healthcare Research and Quality (AHRQ) grant award 5R03HS029417-02. The funders played no role in study design, data collection, analysis and interpretation of data, or the writing of this manuscript.

